# Distinctive trajectories of COVID-19 epidemic by age and gender: a retrospective modeling of the epidemic in South Korea

**DOI:** 10.1101/2020.05.27.20114819

**Authors:** Xinhua Yu, Jiasong Duan, Yu Jiang, Hongmei Zhang

## Abstract

**Objectives:** Elderly people had suffered disproportional burden of COVID-19. We hypothesized that males and females in different age groups might have different epidemic trajectories.

**Methods:** Using publicly available data from South Korea, daily new COVID-19 cases were fitted with generalized additive models, assuming Poisson and negative binomial distributions. Epidemic dynamics by age and gender groups were explored with interactions between smoothed time terms and age and gender.

**Results:** A negative binomial distribution fitted the daily case counts best. Interaction between the dynamic patterns of daily new cases and age groups was statistically significant (p<0.001), but not with gender group. People aged 20-39 years led the epidemic processes in the society with two peaks: one major peak around March 1 and a smaller peak around April 7, 2020. The epidemic process among people aged 60 or above was trailing behind that of younger people with smaller magnitude. After March 15, there was a consistent decline of daily new cases among elderly people, despite large fluctuations of case counts among young adults.

**Conclusions:** Although young people drove the COVID-19 epidemic in the whole society with multiple rebounds, elderly people could still be protected from virus infection after the peak of epidemic.

## Introduction

The novel Severe Acute Respiratory Syndrome associated beta-coronavirus (SARS-CoV-2) of unknown origin, appeared in Wuhan, China in late December 2019 and has swept the world over the past few months (Anderson et al. 2020; Li et al. 2020a; Zhu et al. 2020), causing over 491,500 deaths worldwide (https://coronavirus.jhu.edu/map.html, accessed on June 26, 2020) and significantly disrupting both societal activities and person life(Center 2020). Although several early studies described the dynamics of the epidemic process in details (Li et al. 2020a; Wu and McGoogan 2020), many uncertainties remained. For example, diagnosis criteria varied significantly across countries. During the early epidemic in Wuhan, China, patients were required to have serious pneumonia symptoms plus lab confirmed virus detection (Huang et al. 2020; Zhu et al. 2020), thus missing most mildly symptomatic and all asymptomatic patients. As suggested in a modeling study, probably 86% of COVID-19 cases might be undocumented in Wuhan (Li et al. 2020b). Many epidemic measures such as basic reproduction number based on early epidemic in Wuhan were questioned by later studies due to possible underestimating the true parameters (Nishiura et al. 2020; Zhao et al. 2020a; Zhao et al. 2020b). On the other hand, some countries such as South Korea and Singapore classified patients only based on lab tests, yielding a better picture of the epidemic.

To fully understand the epidemic process of COVID-19, accurate and complete epidemic data are indispensable. Data from South Korea have been generally considered of highest quality, mainly due to two notable strategies adopted by the South Korea government from the beginning of the epidemic: extensive contact tracing and massive testing to identify possible cases in addition to case isolation (Shim et al. 2020). South Korea identified the first COVID-19 case on Jan 20, 2020, and the outbreak started its exponential growth after Feb 19, 2020. In an outbreak occurred in a call center, 1,143 people were tested, 97 were positive and confirmed (positive rate 8.5%) (Park et al. 2020). After tracing all contacts of those 97 cases, about 16% were tested positive (secondary attack rate). In addition, South Korea also installed roadside testing stations to test any person who had concerns about his/her infectious status, in addition to those who had contacted known patients. Such extensive controlling measures not only halted the epidemic successfully but also produced a more complete picture of the COVID-19 epidemic.

A striking phenomenon in COVID-19 was that people aged 65 or older suffered the heaviest burden of the disease (Richardson et al. 2020; Wu and McGoogan 2020) and the proportion of cases was higher in men than that of women. According to a recent CDC report, about 80% of deaths occurred among elderly people, and those aged 80 or above had almost 15% chance of dying if infected (CDC 2020; Garg et al. 2020). In our previous analysis based on Florida COVID-19 data, we found that people aged 65 or older accounted for 54% of hospitalizations and 82% of deaths. The mortality rate was 14% among elderly people who were infected with coronavirus (Yu 2020a).

Furthermore, since May 1, 2020, the COVID-19 pandemic has been waning down across the world (https://coronavirus.jhu.edu/map.html), pressing many countries to consider re-opening the business. Many public health experts warned a possible rebound of new cases if current interventions were relaxed (Chowell and Mizumoto 2020; Ferguson et al. 2020; Kissler et al. 2020). A recent model predicted that COVID-19 epidemic might last more than a year and multiple waves of outbreaks were possible (Kissler et al. 2020). It is likely elderly people may still suffer the heaviest disease burden during the return of outbreak (Hay et al. 2020).

However, it was unknown whether and how the epidemic processes were different between young and old people. In this study, we aim to statistically learn the dynamics of the COVID-19 pandemic based on data from South Korea. In addition to identifying the best fit of the epidemic process, we explore gender- and age group- specific trajectories of COVID-19 to facilitate our understanding of the disease and its impact on different populations, and inform the potential and severity of COVID-19 rebound.

## Materials and Methods

The daily counts of confirmed new COVID-19 cases and deaths were obtained from the open source (https://github.com/jihoo-kim/Data-Science-for-COVID-19, accessed on May 2, 2020), which were systematically gathered from Korea Center for Disease Control (KCDC) daily reports. All cases were verified against KCDC reports. The line list file included patient’s age, gender and date of virus infection confirmation. However, the line list file excluded almost all cases occurred in the city of Daegu (more than 6,000 cases), and thus cases from Daegu were excluded from our study. We further excluded cases with missing confirmation date (n=3). Age was grouped (in years) as 0-19, 20-39, 40-59, and 60 or above. Those with missing gender information (n=78) or missing age information (n=86) were retained in the analysis for overall trajectories (total sample size n=3349), but were excluded in the gender or age specific analysis. Since our purpose in this study was not to predict new cases in the future but to model the epidemic process, we adopted a semi-parametric Generalized Additive Model (GAM) to obtain fitted daily case counts and also account for non-linear patterns of epidemic process (Wood 2017). The time was modeled as a continuous variable with smoothing terms (thin plate regression splines with 8 knots). Interactions between smooth terms and gender (or age group) were modeled as separate smoothing function for each group. Specifically, for interaction models:

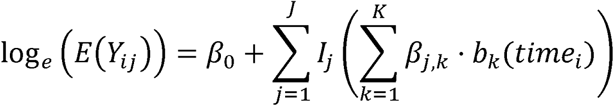

where *Y*_*ij*_ represents the observed case counts of day *i* and group *j* that follows a certain distribution. In this study, we focused on Negative Binomial (NB) or Poisson distributions due to their robustness. We use variable *time*_*i*_ to represent day starting from 1, *I*_*j*_() is an indicator variable (0/1) denoting if daily counts of new cases is for group *j* (1) or not (0), *b*_*k*_() represents a basis function for the *k*^*th*^ term to smooth temporal trend, and *β*_*j,k*_ are regression coefficients for smooth term *k* and group *j* (representing group-specific effects). Parameters were estimated via the restricted maximum likelihood (REML) approach. The Generalized Cross Validation criterion with Mallows’ Cp (GCV.Cp) and Maximum Likelihood (ML) methods were also explored. Therefore, the above GAM framework allows us to compare different trajectories through examining the interactions between smoothed time term and age/gender groups with a focus on comparing the overall trajectories rather than point-wise comparisons. Statistics R^2^ and percent of deviance explained by the models were used to identify the best fit model. R package *mgcv* was used to fit the GAM model (Wood 2017). The data and programs are available online at https://github.com/Jiasong-Duan/COVID-19-epidemic-trajectories.

## Results

From Feb. 19 to Apr. 30, 2020, there were 3,349 COVID-19 cases (1,439 males, 43%) identified outside the Daegu city. Those with age 0-19 accounted for 6% (n=202) of total cases, and age 20-39 for 37% (n=1,227), age 40-59 for 31% (n=1,034), while those with age 60 or above accounted for 24% (n=800) of total cases. As shown in Figure 1, the epidemic outside the Daegu city peaked around Mar. 1, 2020 and declined afterwards except for a second small peak around March 28, 2020.

**Figure 1:**
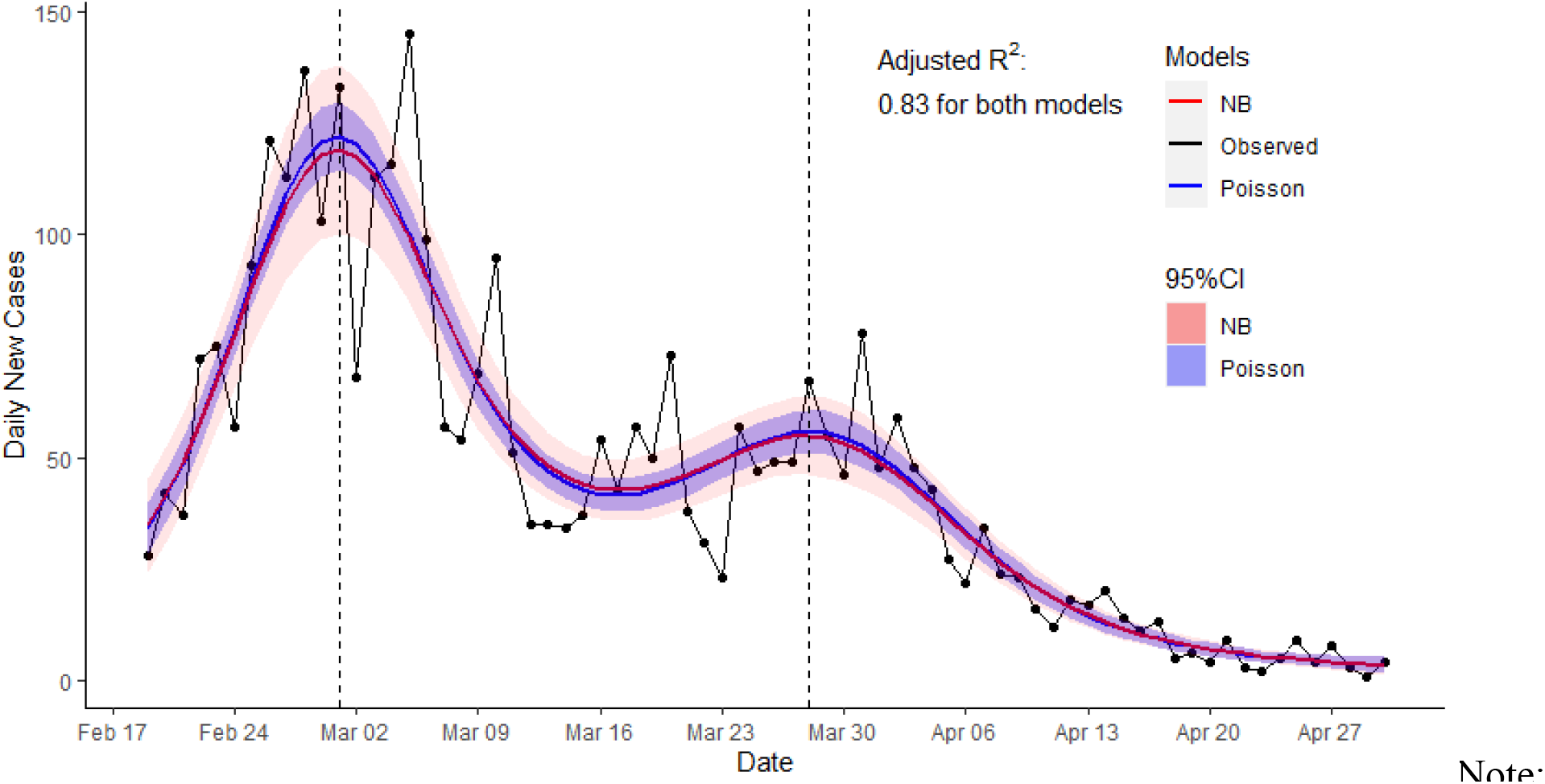
Epidemic curve of COVID-19 and predictions from generalized additive models, South Korea, Feb 19 to April 30, 2020 NB: negative binomial

The fitted curves to the observed daily new cases were overlaid on the observed counts in Figure 1. Predictions from both NB and Poisson models were indistinguishable. However, the confidence intervals from NB model were much wider than that of Poisson model. As shown in the model comparison table (Table 1), there was no difference in the adjusted R^2^ and percent deviance explained by the same model between different estimating methods. The adjusted R^2^ was 0.839 with 89.2% deviance explained by the Poisson model, while the adjusted R^2^ from NB model was 0.838 with 90.3% deviance explained by the model (Table 1). Although both models resulted in similar model fitting parameters, the NB model also estimated a dispersion factor of 18.2, implying Poisson distribution might not be a suitable choice to fit the data. In addition, the wider confidence intervals from the NB model covered a larger range of observed values. Thus, to be conservative, the model based on NB distribution was selected and implemented in the subsequent analyses. The confidence intervals from the fitted models were omitted in the subsequent plots to emphasize different overall patterns in the epidemic process.

**Table 1:** Model comparisons for fitting the COVID-19 epidemic curves, South Korea

| **model** | **method** | **$R^2_{adjusted}$** | **Deviance explained** |
| --- | --- | --- | --- |
| Poisson | REML(default) | 0.8327 | 89.10% |
|  | GCV.Cp | 0.8332 | 89.10% |
|  | ML | 0.8328 | 89.10% |
| NB | REML(default) | 0.8319 | 90.30% |
|  | GCV.Cp | 0.8319 | 90.30% |
|  | ML | 0.8322 | 90.30% |

Note:
REML: REstricted Maximum Likelihood; GCV.Cp: Generalized Cross Validation Criterion with Mallows' Cp; ML: Maximum Likelihood
NB: negative binomial

Figure 2a-b presented the fitted epidemic processes by gender and age groups. The epidemic curve for males fell significantly below that of females (p = 0.0006). Although the epidemic curve of males peaked about one day earlier than that of females as shown in Figure 2a, the shapes of the curve were not significantly different between males and females (p for interaction =0.35). On the other hand, age-specific epidemic curves depicted significantly different patterns across age groups (p for interaction<0.001) (Figure 2b). The epidemic curve in the youngest group (aged 0-19) showed lowest daily case counts and largely stable over the whole period, while there were two peaks in the epidemic process among people aged 20-39 years. In fact, the epidemic among people aged 20-39 led the whole epidemic process in the total population such that not only did young adults have more daily new cases than that of other age groups, but also the epidemic processes among people aged 40-59 and 60+ years were trailing one to three days behind that of aged 20-39.

**Figure 2:**
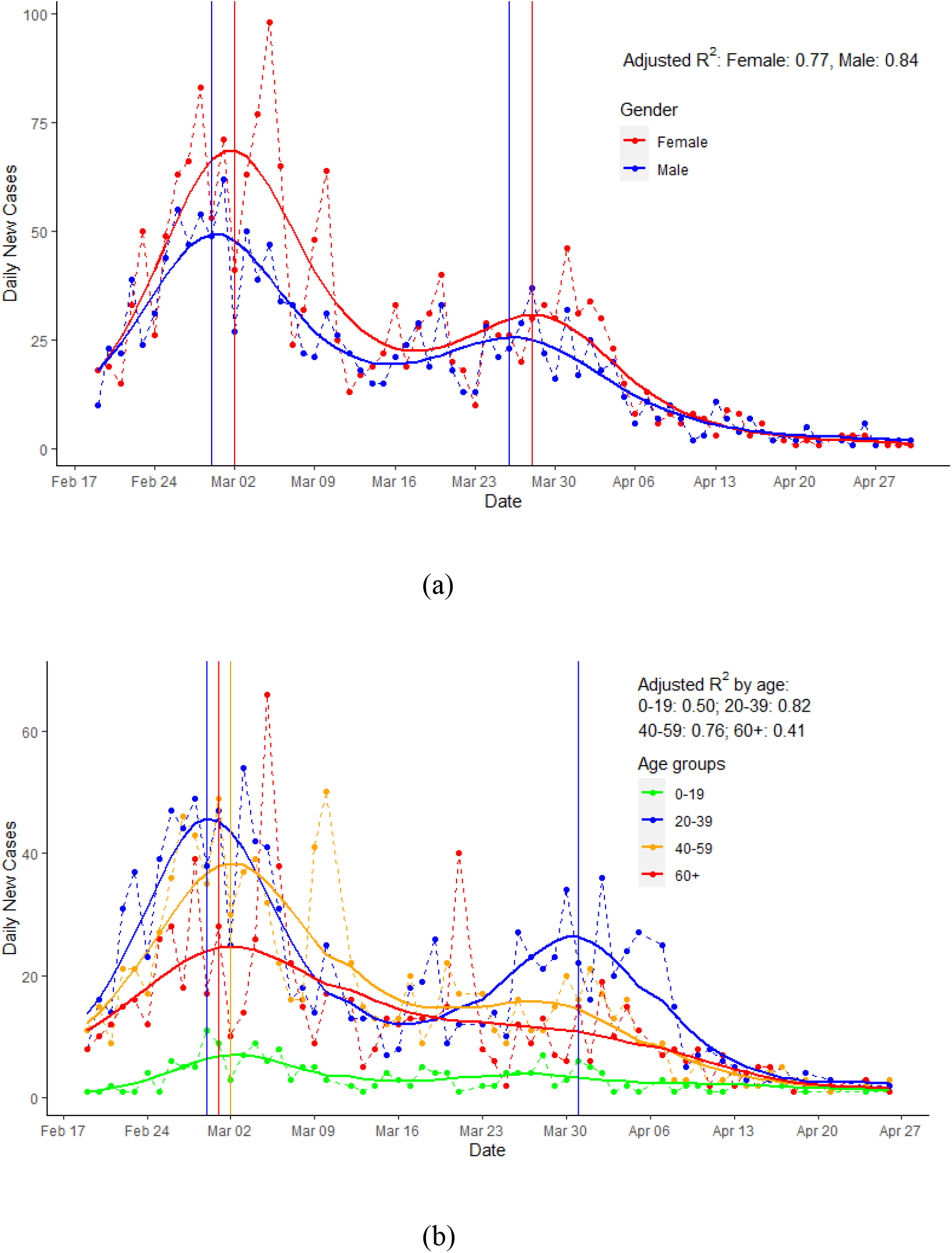
Trajectories of COVID-19 epidemic process by gender(a) and age groups (b), South Korea

To further explore age and gender effects on the epidemic process, Figure 3a-b presented the fitted epidemic curves by age groups for males and females separately. Among males, people aged 20-39 had highest predicted daily counts and experienced two peaks over time, while those aged 60 or older had much lower daily case counts and decreased consistently over time despite the large changes of epidemic in young adults. Those aged 40-59 also experienced two peaks in the epidemic but were at a smaller scale than young adults.

**Figure 3a-b:**
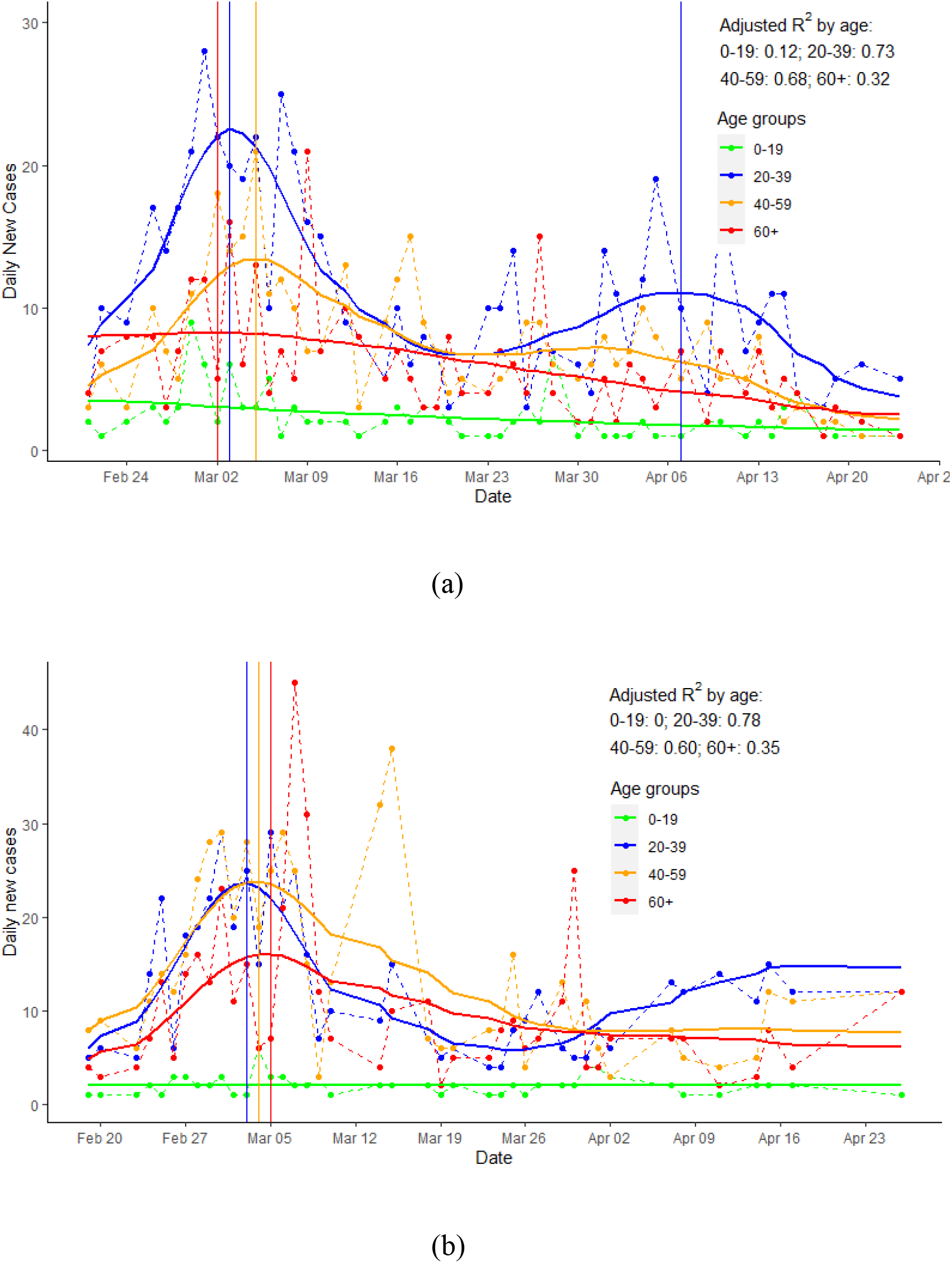
Trajectories of COVID-19 epidemic by age groups, South Korea, for males (a) and females (b) separately

The patterns of epidemic processes by age groups among females were different from that of males. Those females aged 40-59 and aged 20-39 had similar epidemic processes during the first peak of epidemic. The daily case counts among females aged 20-39 also increased after April 1, 2020. Females aged 60 or above had smaller magnitude of epidemic but overall, similar to that of females aged 40-59.

## Discussion

In this study, we demonstrated different trajectories of COVID-19 epidemic between gender and age groups based on South Korea data. First, based on case reporting date and assuming similar incubation periods and reporting delays across all groups and over the whole study period, young people aged 20-39 years led the epidemic processes in the whole society and also had experienced two peaks about one-month apart, one major peak around March 1 and a smaller peak around April 7, 2020; Second, school age people (aged 0-19) had much smaller magnitude of epidemic overall; and finally, the epidemic process among people aged 60 or above was trailing behind that of younger people, and the magnitude of epidemic was smaller than that of people aged 20-39 or 40-59. After March 15, there was a steady decline of daily new cases among people aged 60 or above, despite large fluctuations of case counts among young adults.

Our findings were consistent with other reports in which younger people accounted for most confirmed COVID-19 cases (Guan et al. 2020; Wu and McGoogan 2020; Zhang et al. 2020). Our empirical evidence from high quality data supported that COVID-19 epidemic was driven by the infection among young adults. In addition, school age children had the least burden of disease, possibly due to early school closure and vacation breaks during that period. This pattern was different from that of typical respiratory infection diseases such as seasonal flu in which most cases were school age children.

Worldwide, people aged 60 or above endured a disproportional burden of COVID-19 disease (Wu and McGoogan 2020). They had a higher risk of hospitalizations, and about 80% deaths occurred in this age group(Garg et al. 2020). However, it was unclear whether elderly people were more likely to get infected, whether virus transmissibility was higher among elderly, or whether elderly people were merely more likely to have severe diseases than younger people (Hay et al. 2020; Zhang et al. 2020). Elderly people generally have weaker immune system than younger people. Meanwhile, they have been exposed to many viruses over lifetime that may shield them from getting infected by a new virus, but there was no evidence for any prior immunity to the SARS-CoV2. Nonetheless, our findings provided some hope for mitigating the impact of epidemic on this vulnerable population. As demonstrated in Figure 2b and 3a-b, fitted daily case counts among those aged 60 or older declined consistently after March 15, 2020, despite a second peak occurred in early April among people aged 20-39. Although we did not have detail information about health conditions and behavioral changes among South Korea elderly people during the COVID-19 pandemic, we believe by promptly isolating cases, extensive contact tracing and quarantine at risk people early and efficiently, together with social distancing, avoiding contacting with young cases and proper personal protection (Anderson et al. 2020; Shim et al. 2020), elderly people could be effectively protected from virus infection despite a second rebound in young adults. South Korea had set an excellent model for other country to consider. For example, there were only about ten thousand cases and 282 deaths so far during the COVID-19 epidemic in South Korea (http://www.cdc.go.kr/cdc_eng/, accessed on June 25,2020).

In addition, although overall gender difference in the COVID-19 epidemic was moderate, age and gender specific analyses suggested that females (and to a less extent, males) aged 40-59 had similar experience of epidemic to that of people aged 20-39. This might be because this age group often had close and frequent contacts with younger people in work or within households. Though the risks of hospitalizations and deaths were low among this population, they were higher than that of regular respiratory infectious diseases such as seasonal flu. Thus, the disease burden among this middle age group should not be neglected.

There were some limitations in this study. First, our study excluded cases from the city of Daegu (over 6000 cases) because detail information about cases from that city was not released to the public. Although it was unlikely to bias our results, information from such a large outbreak could provide some additional insights on how the epidemic unfolded among people of different age and gender. However, during the early stage of epidemic, little gender and age stratified data were publicly available, and most individual level data from other regions were incomplete as well.

Second, we employed statistical methods to examine the trajectories of epidemic. There were two perspectives to model the epidemic process (Hethcote 2000; Unkel et al. 2012). One common approach was to model the process based on the mechanisms of the epidemic. For example, the Susceptible-Exposed-Infectious-Removed (SEIR) model and its variants had been used to assess the dynamic of epidemic, obtain epidemic parameters, and evaluate the impact of various control measures on the epidemic (Kucharski et al. 2020; Peak et al. 2017; Prem et al. 2020; Yu 2020b). Agent-based models were also used to simulate the epidemic process and assess the effects of various interventions (Ferguson et al. 2020; Wu et al. 2020). The other perspective was based on traditional statistical models. Non-linear models such as generalized logistic growth model (Chowell 2017) were used to model the growth of the epidemic and estimate the growth rate of cases over time. In addition, some researchers directly modeled the epidemic curve with regression techniques, assuming daily counts follow some distributions such as Poisson or negative binomial distributions. For example, models based on time series of count data were adopted to predict the COVID-19 deaths in the US, such as those models from Institute of Health Metrics and Evaluation (IHME) (IHME 2020) and University of Texas-Austin (Woody et al. 2020). Our previous research also used vector autoregressive models to examine the risk interactions across age groups after the peak of COVID-19 Epidemic (Yu 2020c). While there were many uncertainties among different gender and age groups about contact patterns, virus transmissibility and behavioral changes during the epidemic, since the epidemic data from South Korea were more likely to be complete, it is possible to directly model the daily counts with regression models assuming a common distribution for count data. We believed that out models avoided many unfounded assumptions in the more complicated epidemic process models.

Third, we only have case reporting or lab confirmation dates in this study which were likely 3-5 days away from the actual virus infection date. The average incubation date for COVID-19 was about 5 days (Lauer et al. 2020) and the report delay in South Korea was unknown but likely very short due to extensive testing. Thus, we make some untestable assumptions in comparing epidemic trajectories between age and gender groups. The incubation period and reporting delay were assumed to be the same across all groups and over the whole study period. This should be pertinent in South Korea as they started mass testing and contact tracing from the beginning of the epidemic (Shim et al. 2020) but may not be appropriate for the regions that testing is severely limited and delayed.

Finally, we only analyzed data from South Korea. The epidemic processes of COVID-19 in different countries were likely different due to different population structure and different interventions to mitigate the epidemic (Anderson et al. 2020; Chowell and Mizumoto 2020; Hay et al. 2020; Lipsitch et al. 2020). Meanwhile, we expect our findings provided a general picture of the epidemic trajectories of COVID-19 and can serve as a reference to other regions. In addition, as witnessed in the COVID-19 epidemic, politics and ideology often overtook science and public health, so that effective interventions were sometimes implemented too late and incomplete, leaving the public at lost and public health practitioners in conundrum.

The main strength of our study was our straightforward analyses to explore different epidemic processes based on high quality data. Insights often emerge through such modeling exercise. We stratified the models by age and gender groups and discovered their different trajectories in the epidemic. Recent studies had predicted a long-lasting epidemic for COVID-19 and possible multiple waves of outbreaks after societal re-opening(Kissler et al. 2020). Our findings were unique in providing empirical evidence for designing effective public health strategies to mitigate the impact of recurrent COVID-19 epidemics and protect vulnerable populations.

## Data Availability

publicly available data

## Conclusions

In summary, in South Korea, and likely in other countries, COVID-19 epidemic processes had distinctive dynamic patterns among age and gender groups. Epidemic among young adults led the epidemic process in the whole population, and a second peak occurred in people aged 20-39 years. More importantly, during the post-peak period of the COVID-19 epidemic and in the process of gradually returning the society and economy to normalcy, elderly people could be protected effectively though case isolation, contact tracing, mass testing, and proper personal protections, as exemplified in South Korea.

## Ethics statement

This study used only publicly available data and no human subjects were directly involved, thus deemed to be exempted from the approval of Institutional Review Board. No informed consent was needed. All authors declared no conflict of interest in conducting this study.

